# A Contribution to the Mathematical Modeling of the Corona/COVID-19 Pandemic

**DOI:** 10.1101/2020.04.01.20050229

**Authors:** Günter Bärwolff

## Abstract

The responsible estimation of parameters is a main issue of mathematical pandemic models. Especially a good choice of *β* as the number of others that one infected person encounters per unit time (per day) influences the adequateness of the results of the model. For the example of the actual COVID-19 pandemic some aspects of the parameter choice will be discussed. Because of the incompatibility of the data of the Johns-Hopkins-University [3] to the data of the German Robert-Koch-Institut we use the COVID-19 data of the European Centre for Disease Prevention and Control [2] (ECDC) as a base for the parameter estimation. Two different mathematical methods for the data analysis will be discussed in this paper and possible sources of trouble will be shown.

Parameters for several countries like UK, USA, Italy, Spain, Germany and China will be estimated and used in W. O. Kermack and A. G. McKendrick’s SIR model[1]. Strategies for the commencing and ending of social and economic shutdown measures are discussed.

The numerical solution of the ordinary differential equation system of the modified SIR model is being done with a Runge-Kutta integration method of fourth order [4].

At the end the applicability of the SIR model could be shown. Suggestions about appropriate points in time at which to commence with lockdown measures based on the acceleration rate of infections conclude the paper. This paper is an improved sequel of [5].

## 1 The mathematical SIR model

At first we will describe the model. *I* denotes the infected people, *S* stands for the susceptible and *R* denotes the recovered people. The dynamics of infections and recoveries can be approximated by the ODE system

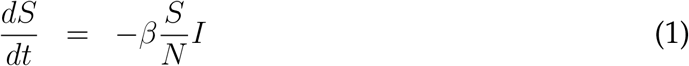

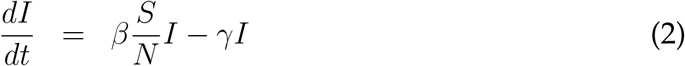

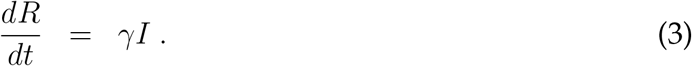

We understand *β* as the number of others that one infected person encounters per unit time (per day). *γ* is the reciprocal value of the typical time from infection to recovery. *N* is the total number of people involved in the epidemic disease and there is *N* = *S* + *I* + *R*.

The empirical data currently available suggests that the corona infection typically lasts for some 14 days. This means *γ* = 1*/*14 ≈ 0,07.

The choice of *β* is more complicated and will be considered in the next section.

## 2 The estimation of *β* based on real data

We use the European Centre for Disease Prevention and Control [2] as a data for the COVID-19 infected people for the period from December 31st 2019 to April 8th 2020.

At the beginning of the pandemic the quotient *S/N* is nearly equal to 1. Also, at the early stage no-one has yet recovered. Thus we can describe the early regime by the equation

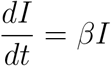

with the solution

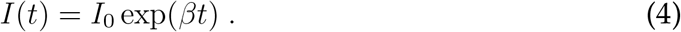

We are looking for periods in the spreadsheets of infected people per day where the course can be described by a function of type (4). Starting with a spreadsheet like

| day | number of infected people |
| --- | --- |
| $t_1$ | $I_1$ |
| $t_2$ | $I_2$ |
| ⋮ | ⋮ |
| $t_k$ | $I_k$ |

for a certain country and a chosen period [*t*_1_, *t*_*k*_] with my favored method We search for the minimum of the functional

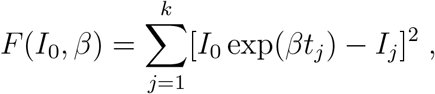

i.e.

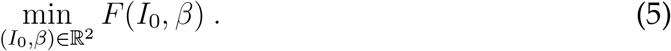

We solved this non-linear minimum problem with the damped Gauss-Newton method (see [4]). After some numerical tests we found the subsequent results for the considered countries. Thereby we chose different periods for the countries with the aim to approximate the infection course in a good quality. The following figures show the graphs and the evaluated parameter.

It must be said that evaluated *β*-values are related to the stated period. For the iterative Gauss-Newton method we guessed the respective periods for every country by a visual inspection of the graphs of the infected people over days.

Especially in medicine, psychology and other life sciences the logarithm behavior of data was readily considered.

Instead of the above table of values the following logarithmic one was used.

| day | log(number of infected people) |
| --- | --- |
| $t_1$ | $\log I_1$ |
| $t_2$ | $\log I_2$ |
| ⋮ | ⋮ |
| $t_k$ | $\log I_k$ |

The logarithm of (4) leads to

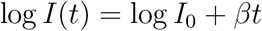

and based on the logarithmic table the functional

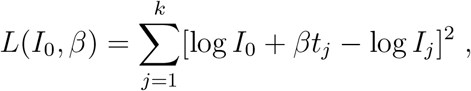

is to minimize. The solution of this linear optimization problem is trivial and it is available in most of computer algebra systems as a “block box” of the logarithmic-linear regression.

The following figures show the results for the same periods as above for Spain, the UK, the USA and Italy.

Figures 7-14 show that the logarithmic-linear regression implies poor results. Thus, the non-linear optimization problem (5) is to choose as the favored method for the estimation of *I*_0_ and *β*. We found some notes on the parameters of Italy in the literature, for example *β* = 0 25, and we are afraid that this is a result of the logarithmic-linear regression.

**Figure 1:**
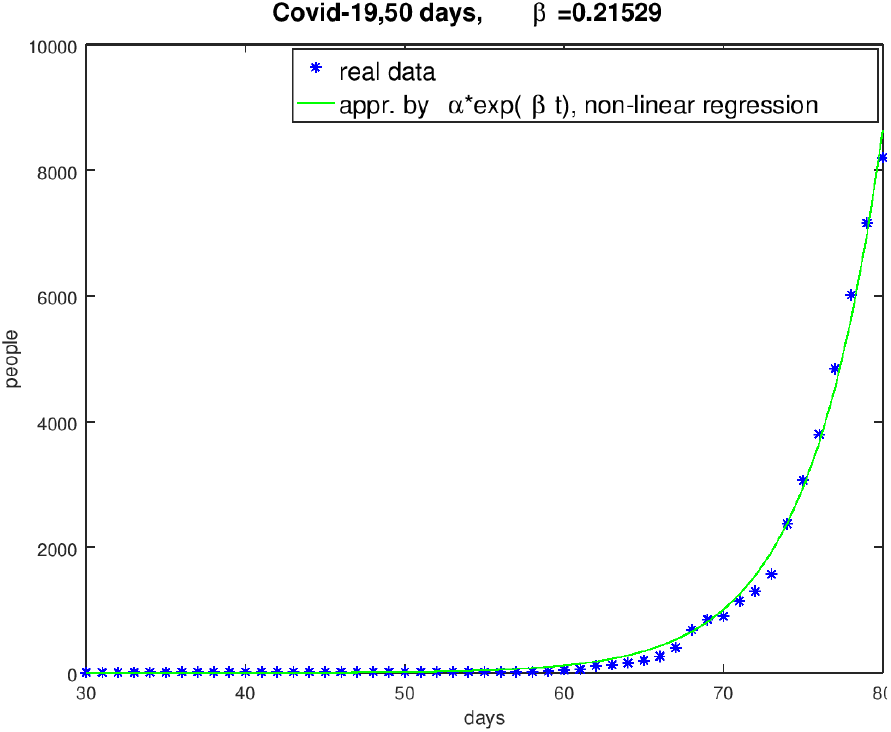
German course from January 31st 2020 to March 20th 2020

**Figure 2:**
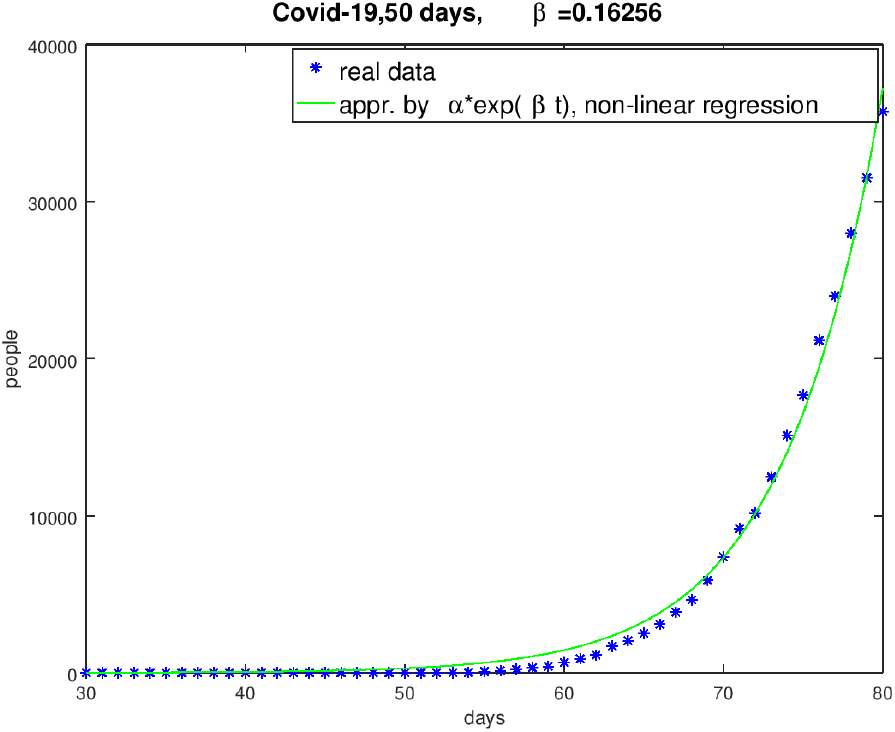
Italian course from January 31st 2020 to March 20th 2020

**Figure 3:**
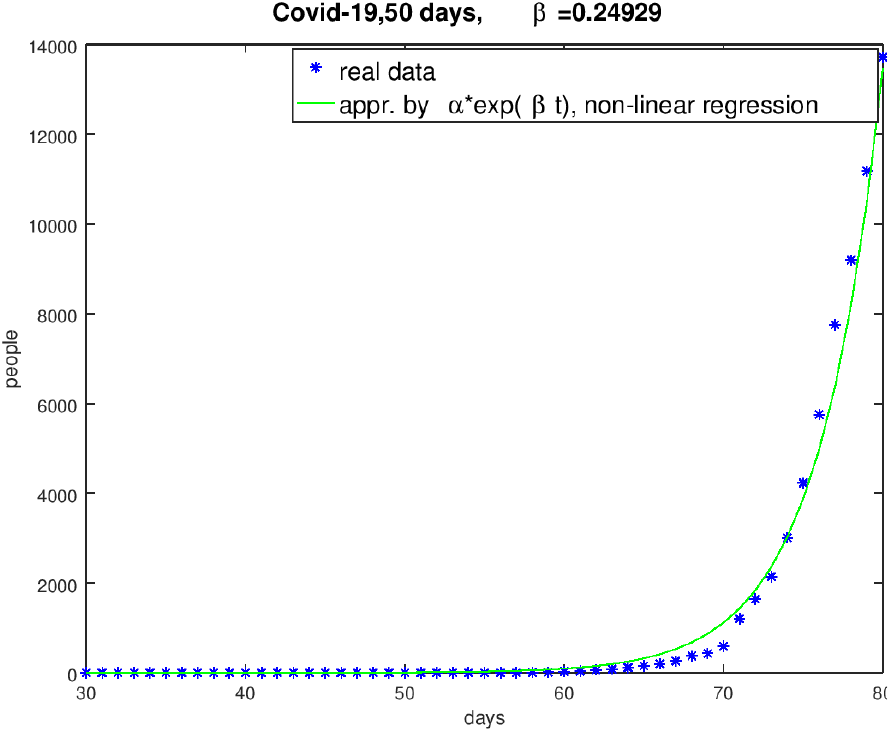
Spanish course from January 31st 2020 to March 20th 2020

**Figure 4:**
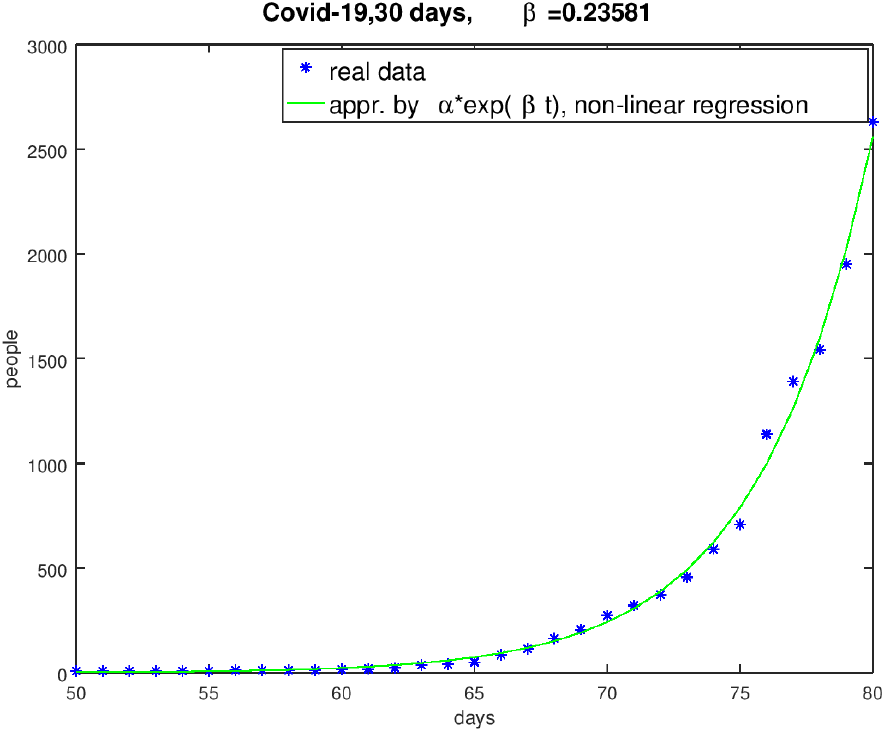
UK course from February 20th 2020 to March 20th 2020

**Figure 5:**
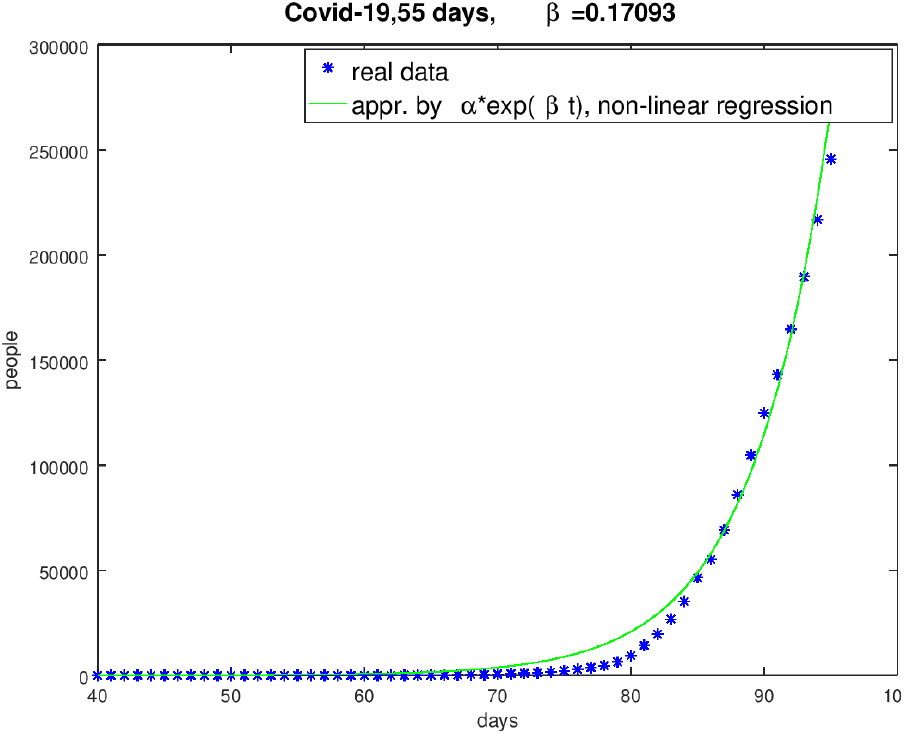
USA course from February 10th 2020 to April 4th 2020

**Figure 6:**
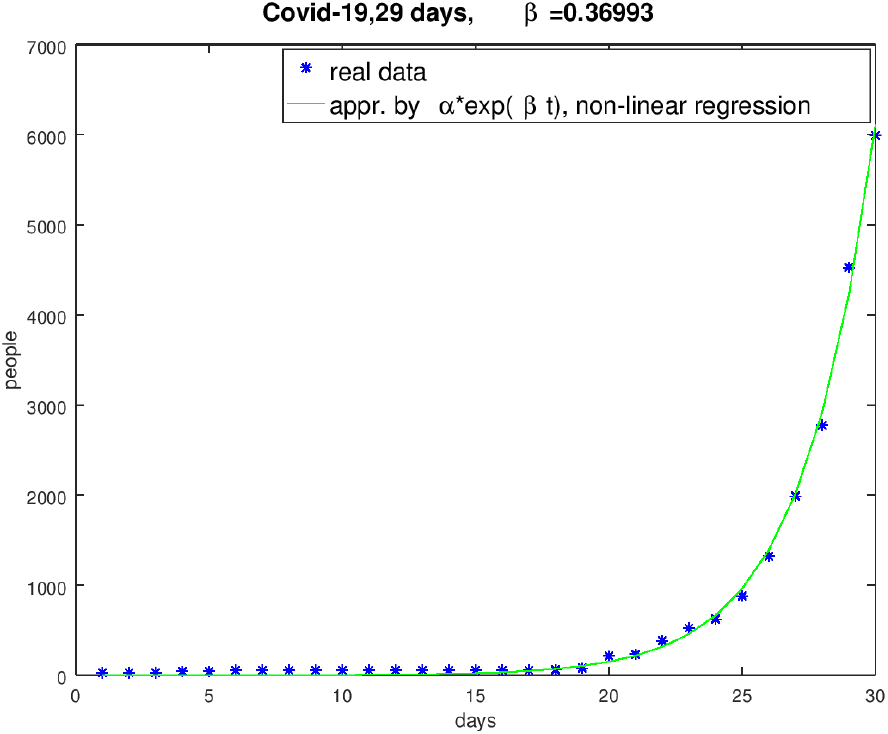
Chines course from December 31st 2019 to January 28th 2020

**Figure 7:**
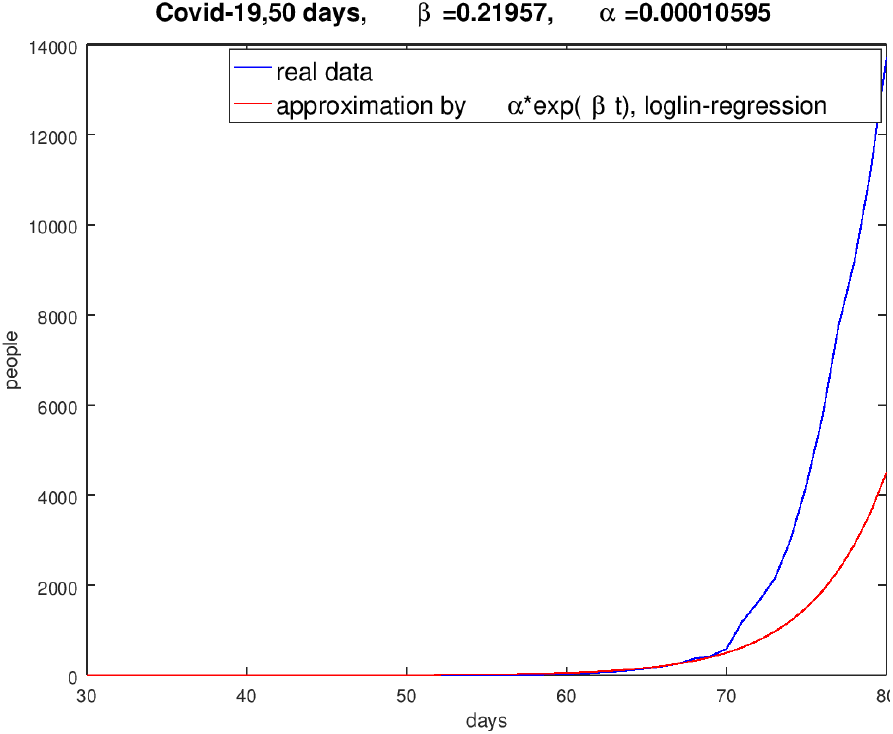
log-lin-result for Spain (January 31st 2020 to March 20th 2020)

**Figure 8:**
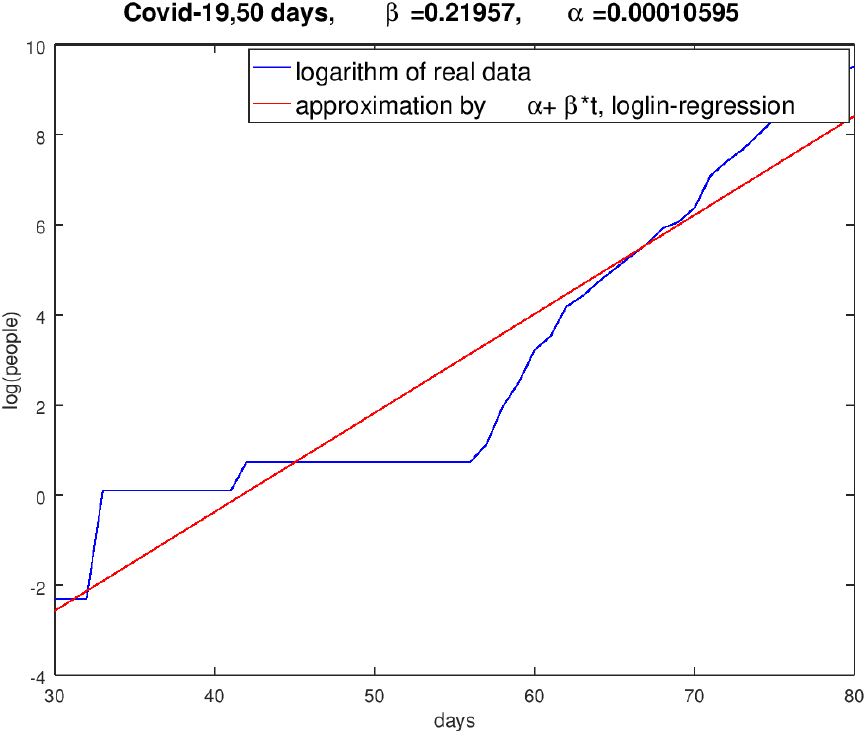
Logarithm of the Spanish result (January 31st 2020 to March 20th 2020)

**Figure 9:**
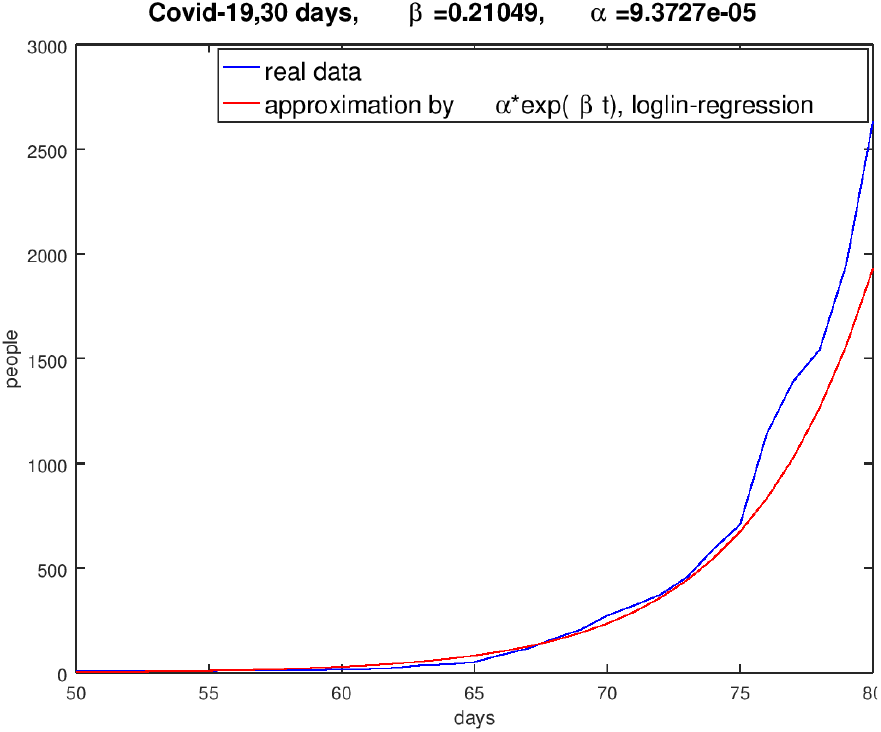
log-lin-result of the UK (February 20th 2020 to March 20th 2020)

**Figure 10:**
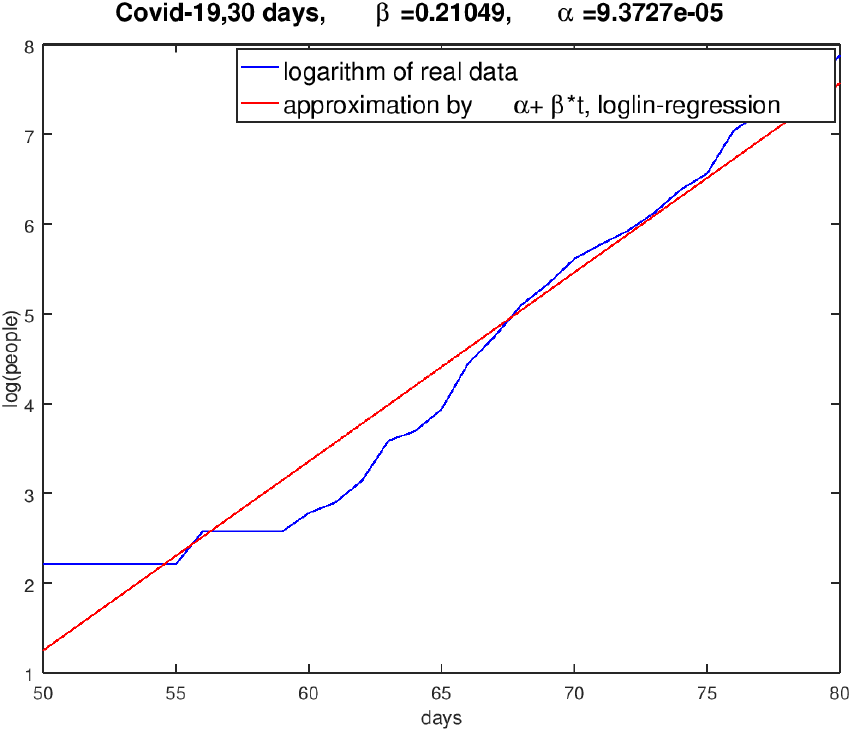
Logarithm of the UK result (February 20th 2020 to March 20th 2020)

**Figure 11:**
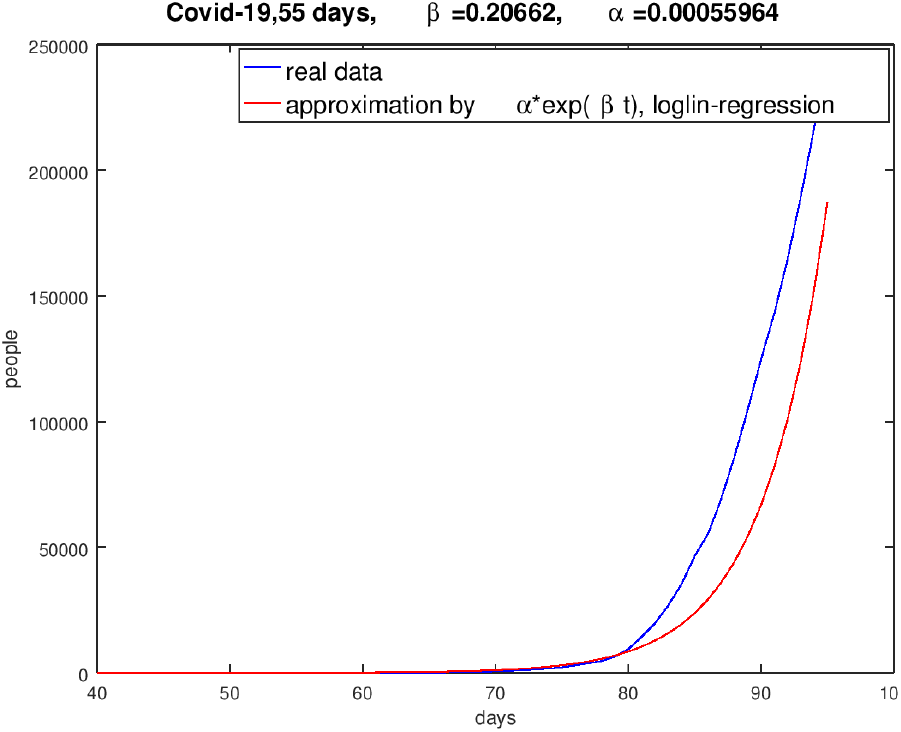
log-lin-result of the USA (February 10th 2020 to April 4th 2020)

**Figure 12:**
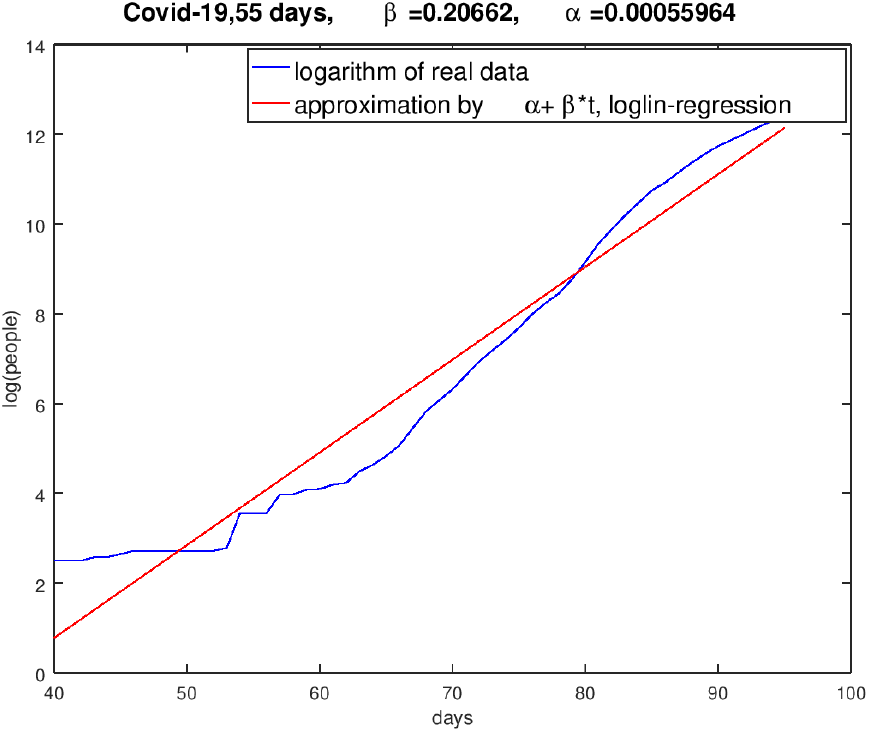
Logarithm of the USA result (February 10th 2020 to April 4th 2020)

**Figure 13:**
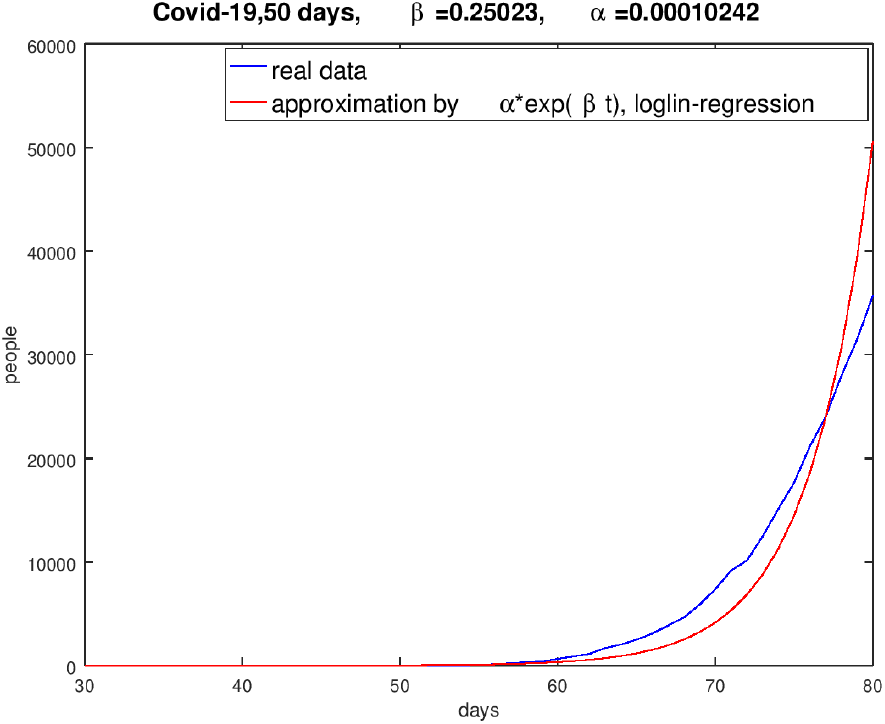
log-lin-result of Italy (January 31st 2020 to March 20th 2020)

**Figure 14:**
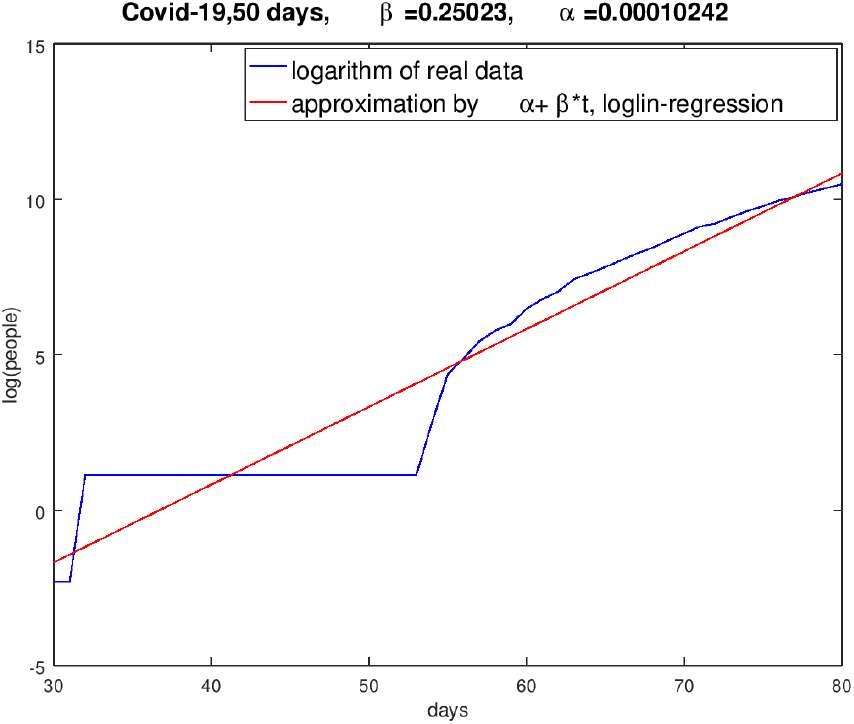
Logarithm of the Italian result (January 31st 2020 to March 20th 2020)

## 3 Some numerical computations for Germany and Spain

With the choice of *β*-value 0,215 (see fig. 1) which was evaluated on the basis of the real data of ECDC and *γ* = 0,07 one gets the course of the pandemic dynamics pictured in fig. 15.^1^. *R*_0_ is the basis reproduction number of persons, infected by the transmission of a pathogen from one infected person during the infectious time (*R*_0_ = *β/γ*) in the following figures.

**Figure 15:**
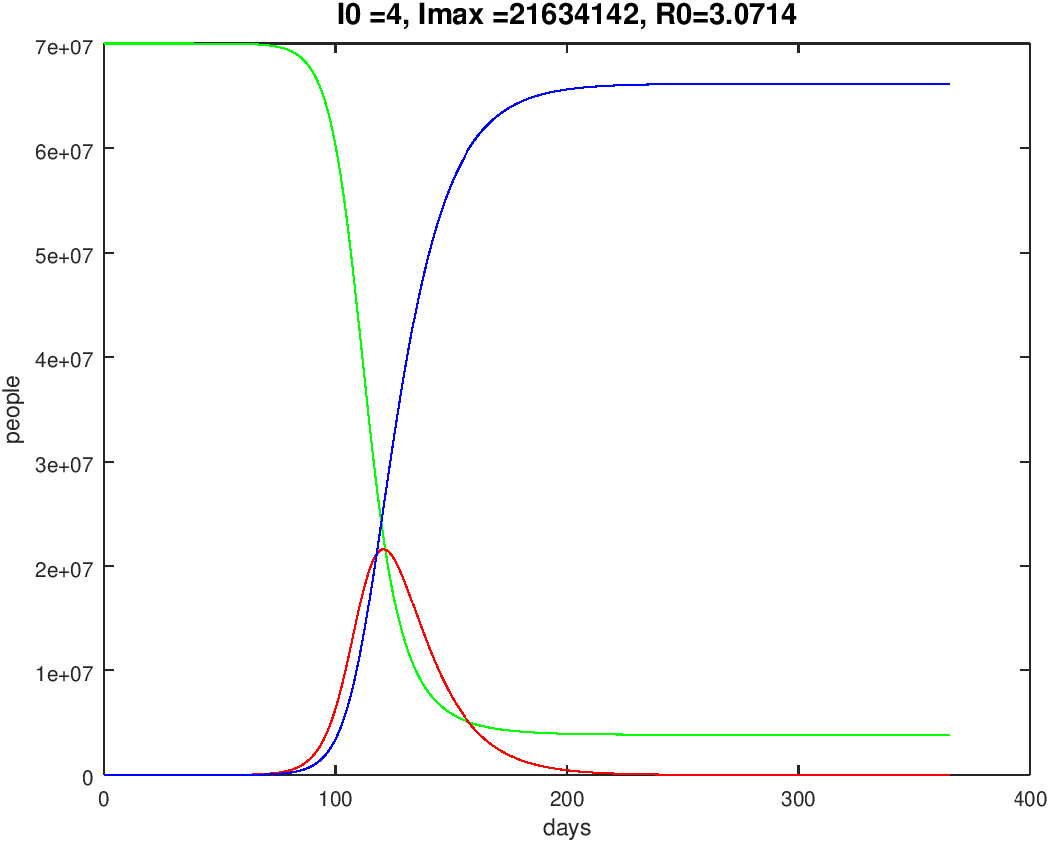
German course of one year, starting end of January 2020, *S*-green, *I*-red, *R*-blue

Neither data from ECDC nor the data from the German Robert-Koch-Institut and the data from the Johns Hopkins University are correct, for we have to reasonably assume that there are a number of unknown cases. It is guessed that the data covers only 15% of the real cases. Considering this we get a slightly changed results and in the subsequent computations we will include estimated number of unknown cases to the initial values of *I*.

For Spain we use the *β*-value 0,249 (see fig. 2) and *γ* = 0,07 we get the course pictured in fig. 16. *N* was set to 40 millions.

**Figure 16:**
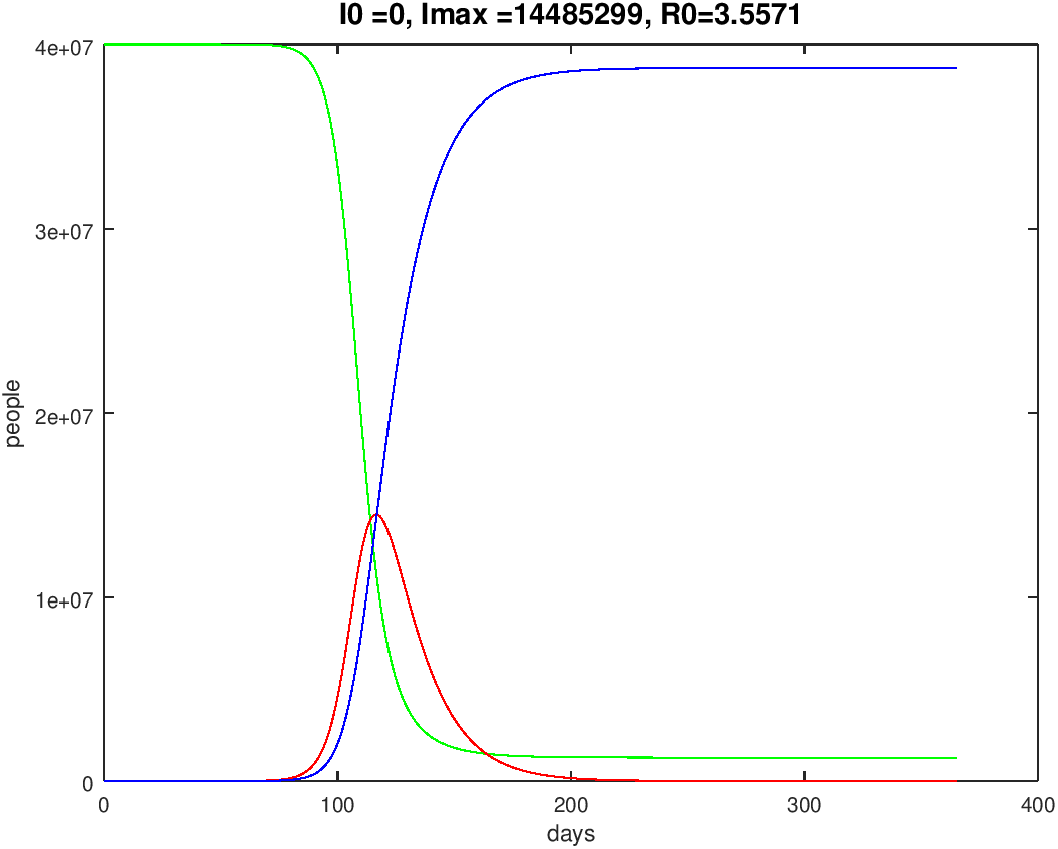
Spanish course of one year, starting end of January 2020, *S*-green, *I*-red, *R*-blue

## 4 Influence of a temporary lockdown and extensive social distancing

In all countries concerned by the Corona pandemic a lockdown of the social life is discussed. In Germany the lockdown started at March 16th 2020. The effects of social distancing to decrease the infection rate can be modeled by a modification of the SIR model. The original ODE system (1)-(3) was modified to

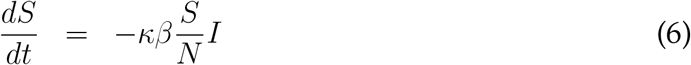

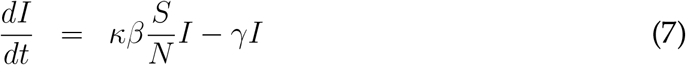

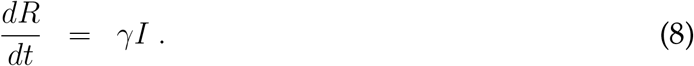

*κ* is a function with values in [0,1]. For example

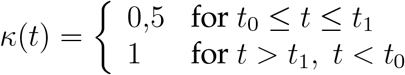

means for example a reduction of the infection rate of 50% in the period [*t*_0_, *t*_1_] (Δ_*t*_ = *t*_1_ − *t*_0_ is the duration of the temporary lockdown in days). A good choice of *t*_0_ and *t*_*k*_ is going to be complicated.

If we respect the chosen starting day of the German lockdown, March 16th 2020 (this conforms the 46th day of the concerned year), and we work with

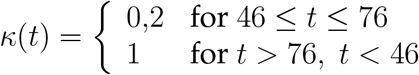

we got the result pictured in fig. 17.

**Figure 17:**
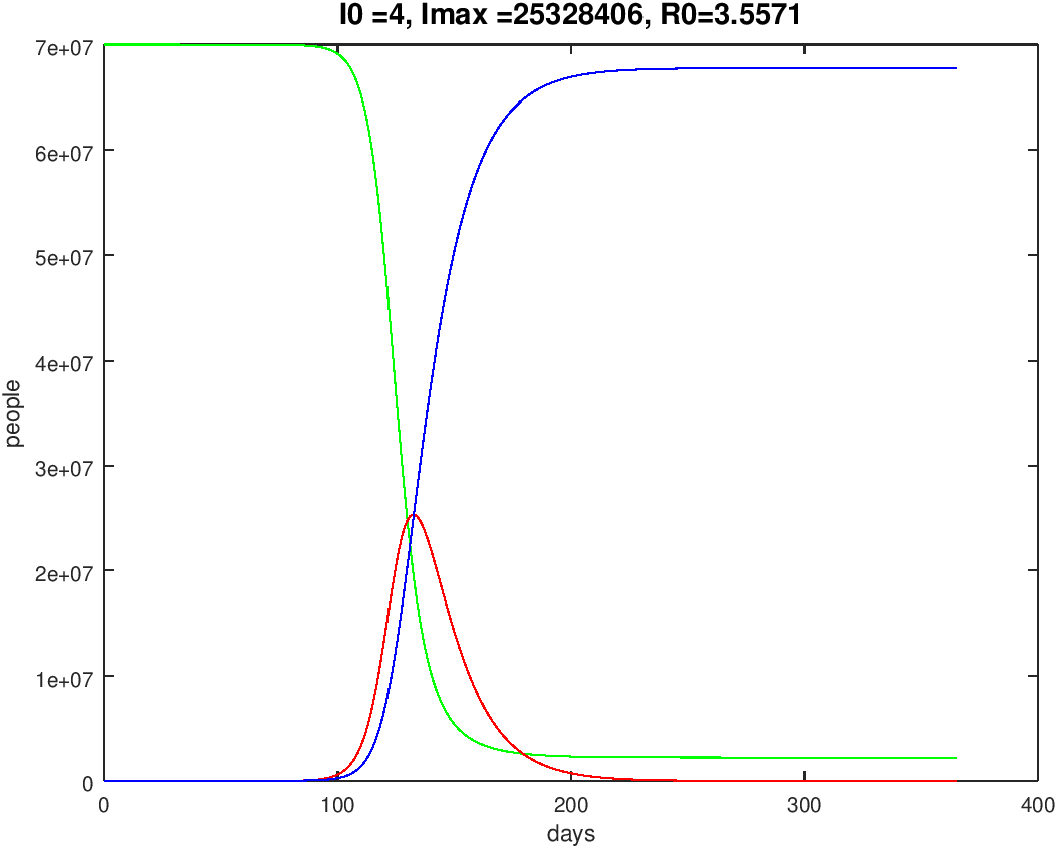
German course of one year, starting end of January 2020, *S*-green, *I*-red, *R*-blue, 30 days lockdown, starting time March 16th 2020

The numerical tests showed that a very early start of the lockdown resulting in a reduction of the infection rate *β* results in the typical Gaussian curve to be delayed by *I*; however, the amplitude (maximum value of *I*) doesn’t really change.

One knows that the development of the infected people looks like a Gaussian curve. The interesting points in time are those where the acceleration of the numbers of infected people increases or decreases, respectively.

These are the points in time where the curve of *I* was changing from a convex to a concave behavior or vice versa. The convexity or concavity can be controlled by the second derivative of *I*(*t*).

Let us consider equation (2). By differentiation of (2) and the use of (1) we get

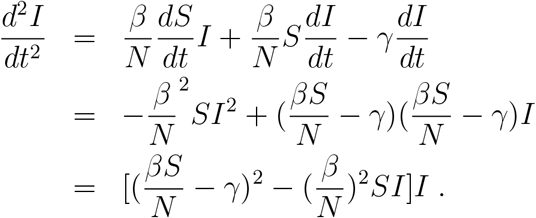

With that the *I*-curve will change from convex to concave if the relation

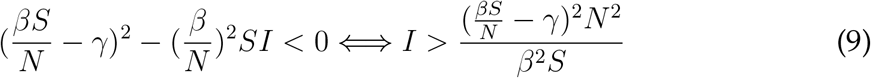

is valid. For the switching time follows

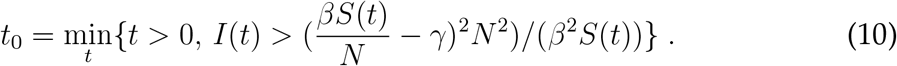

A lockdown starting at *t*_0_ (assigning *β*^*∗*^ = *κβ, κ*∈ [0,1[) up to a point in time *t*_1_ = *t*_0_ + Δ_*t*_, with Δ_*t*_ as the duration of the lockdown in days, will be denoted as a **dynamical lockdown** (for *t* > *t*_1_ *β*^*∗*^ was reset to the original value *β*).

*t*_0_ means the point in time up to which the growth rate increases and from which on it decreases. Fig. 18 shows the result of such a computation of a dynamical lockdown. We got *t*_0_ = 108 (*κ* = 0,2)-The result is significant. In fig. 20 a typical behavior of 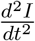 is plotted.

**Figure 18:**
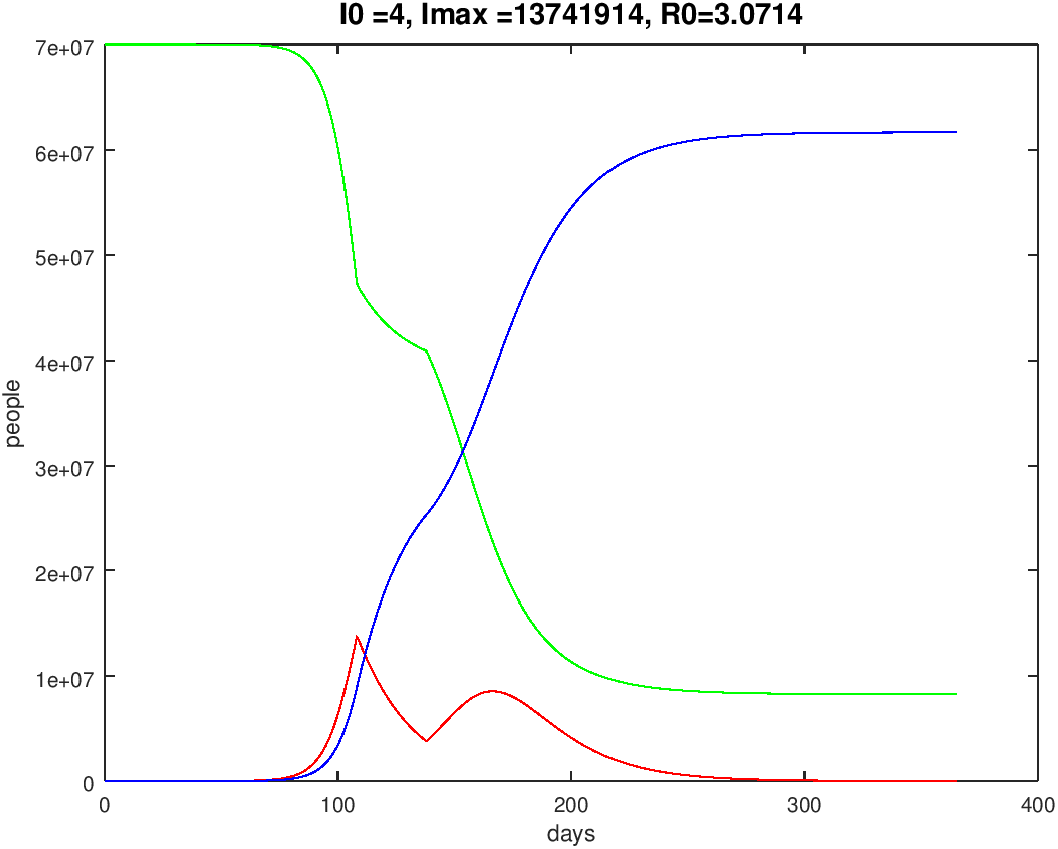
German course of one year, starting end of January 2020, dynamical lockdown, *S*-green, *I*-red, *R*-blue

The result of a dynamical 30 days lockdown for Spain is shown in fig. 19, where we found *t*_0_ = 106 (*κ* = 0,2).

**Figure 19:**
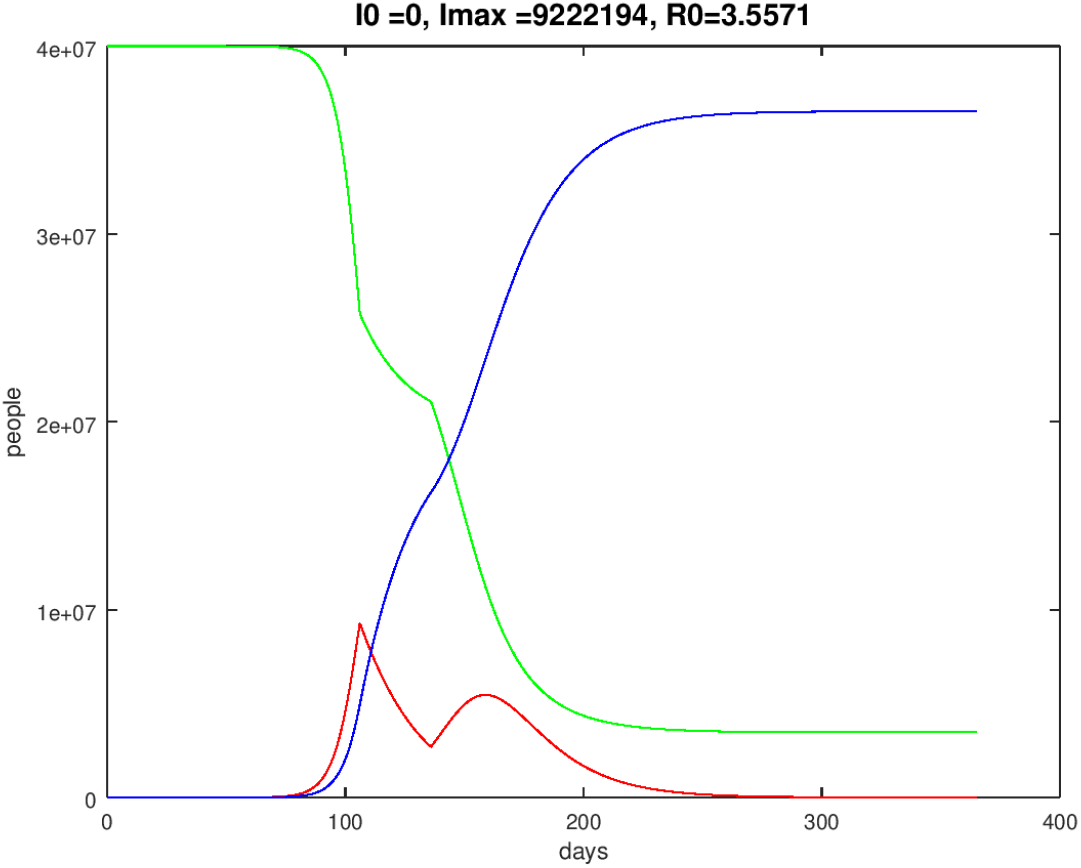
Spanish course of one year, starting end of March 2020, dynamical lockdown, *S*-green, *I*-red, *R*-blue

Data from China and South Korea suggests that the group of infected people with an age of 70 or more is of magnitude 10%. This group has a significant higher mortality rate than the rest of the infected people. Thus we can presume that *α*=10% of *I* must be especially sheltered and possibly medicated very intensively as a highrisk group.

Fig. 21 shows the German time history of the above defined high-risk group with a dynamical lockdown with *κ* = 0,2 compared to regime without social distancing. The maximum number of infected people decreases from approximately 1,7 millions of people to 0,8 millions in the case of the lockdown (30 days lockdown).

**Figure 20:**
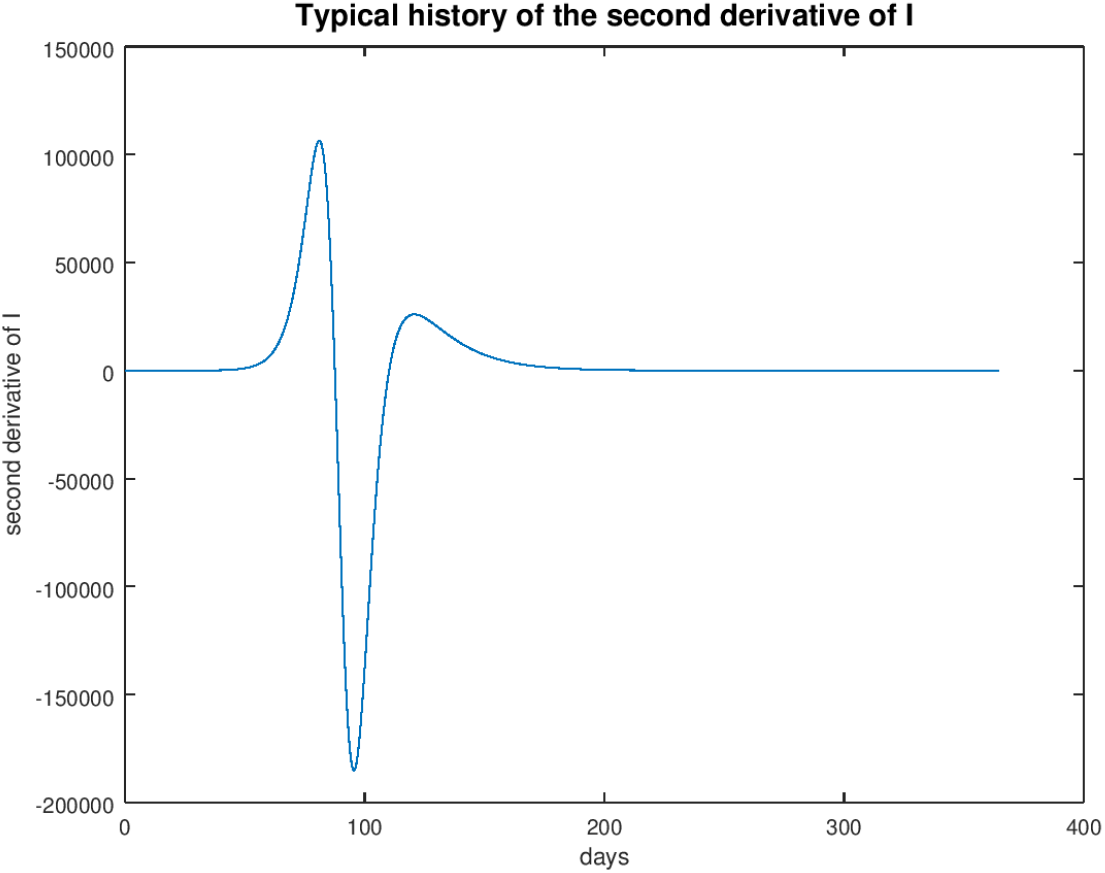
History of the second derivative of *I* (de)

**Figure 21:**
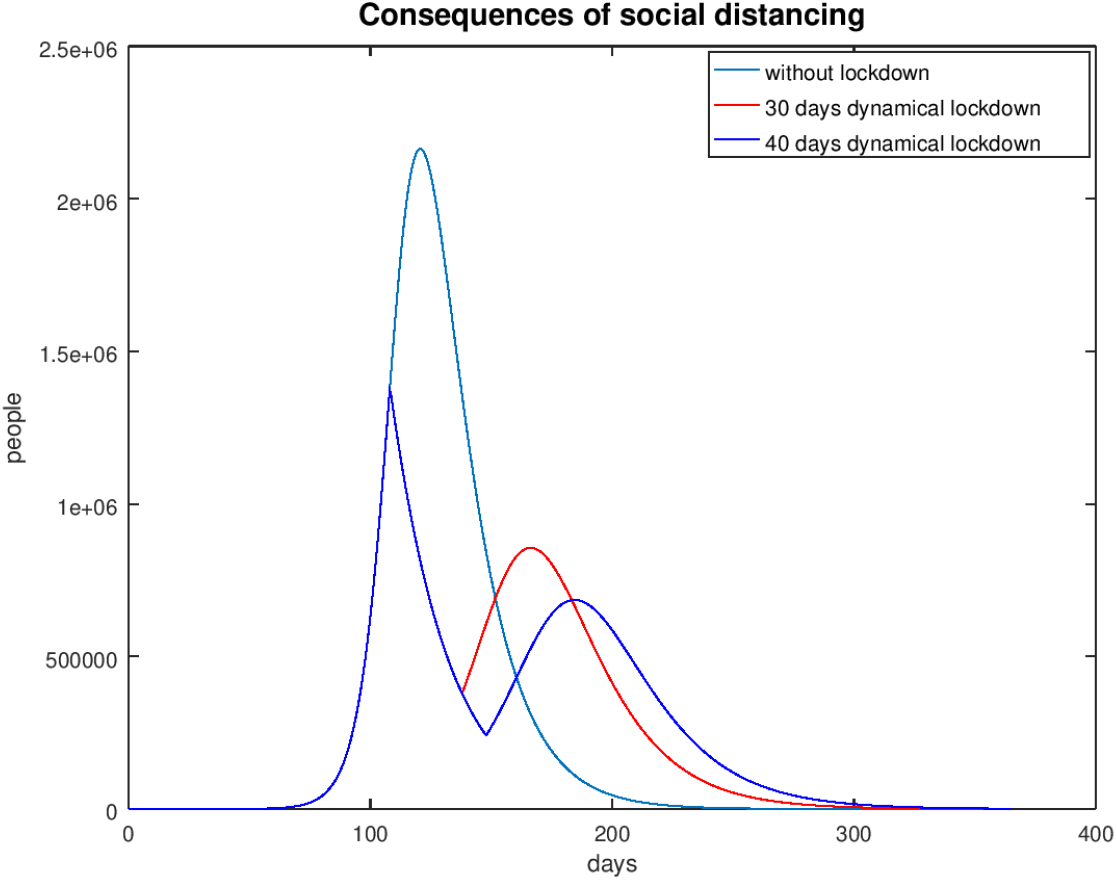
German history of the infected people of high-risk groups depending on a dynamical lockdown

This result proves the usefulness of a lockdown or a strict social distancing during an epidemic disease. We observe a flattening of the infection curve as requested by politicians and health professionals. With a strict social distancing for a limited time one can save time to find vaccines and time to improve the possibilities to help high-risk people in hospitals.

To see the influence of a social distancing we look at the spanish situation without a lockdown and a dynamical lockdown of 30 days with fig. 22 (*κ* = 0,2) for the 10% high-risk people.

**Figure 22:**
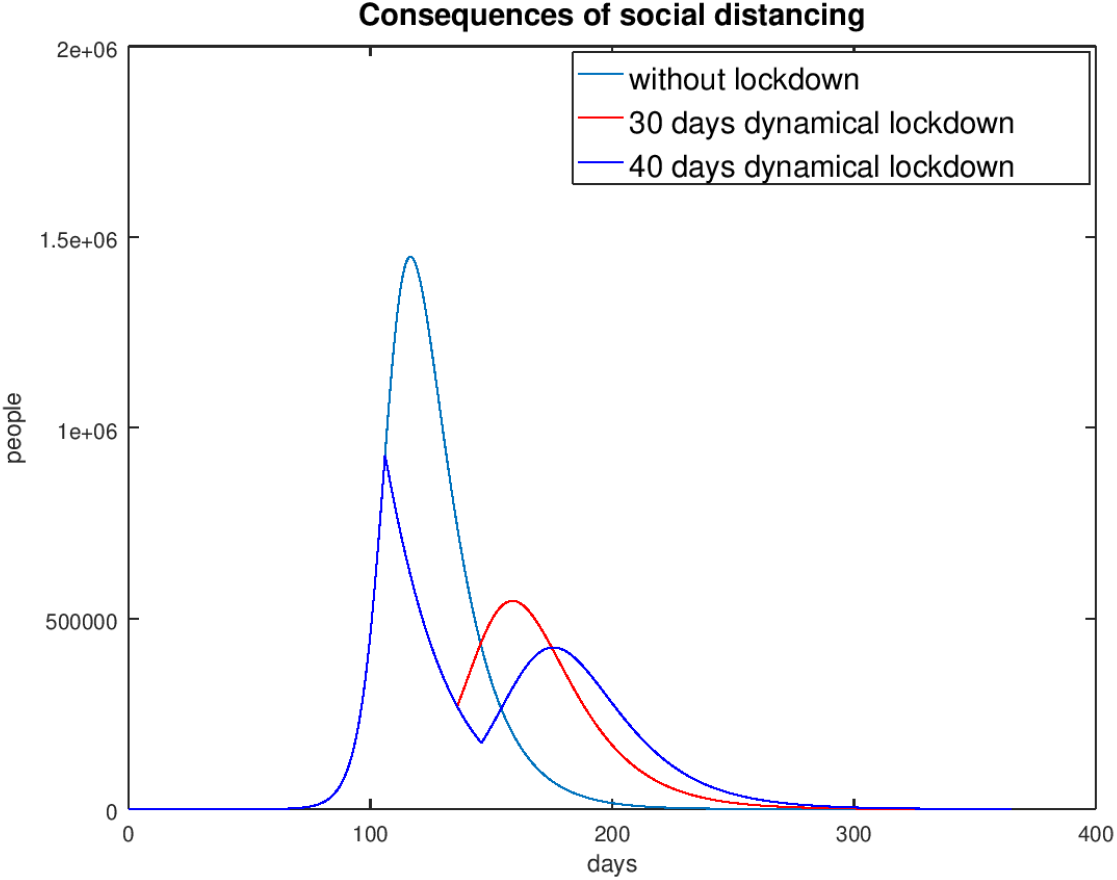
Spanish history of the infected people of high-risk groups depending on a dynamical lockdown

The computations with the SIR model show, that the social distancing with a lockdown will only be successful with a start behind the time greater or equal to *t*_0_, found by the evaluation of the second derivative of *I* (formula (10)). If the lockdown is started at a time less then *t*_0_ the effect of such a social distancing is not significant.

## 5 Closing remarks

If we write (2) or (7) resp. in the form

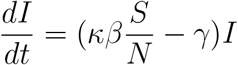

we realize that the number of infected people decreases if

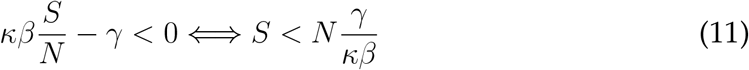

is complied. The relation (11) shows that there are two possibilities for the rise of infected people to be inverted and the medical burden to be reduced.

a. The reduction of the stock of the species *S*. This can be obtained by immunization or vaccination. Another possibility is the isolation of high-risk people (70 years and older). Positive tests for antibodies reduce the stock of susceptible persons.
b. A second possibility is the reduction of the infection rate *κβ*. This can be achieved by strict lockdowns, social distancing at appropriate times, or rigid sanitarian moves.

The results are pessimistic in total with respect to a successful fight against the COVID-19-virus. Hopefully the reality is a bit more merciful than the mathematical model. But we rather err on the pessimistic side and be surprised by more benign developments.

Note again that the parameters *β* and *κ* are guessed very roughly. Also, the percentage *α* of the group of high-risk people is possibly overestimated. Depending on the capabilities and performance of the health system of the respective countries, those parameters may look different. The interpretation of *κ* as a random variable is thinkable, too.

## Data Availability

I use actual data if coronavirus infected people given by the Johns-Hopkins-University

https://www.jhu.edu

## Footnotes

1 *I*0 denotes the initial value of the *I* species, that is January 31th 2020. *Imax* stands for the maximum of *I*. The total number *N* for Germany is guessed to be 75 millions.

